# Comparing Strategies to Introduce Two New Antibiotics for Gonorrhea: A Modeling Study

**DOI:** 10.1101/2025.07.01.25330638

**Authors:** Madeleine C. Kline, Kirstin Oliveira Roster, David Helekal, Eva Rumpler, Yonatan H. Grad

## Abstract

**Introduction:** Drug resistance in *Neisseria gonorrhoeae* is an urgent public health threat. The anticipated approval of two new antimicrobials for gonorrhea prompts the need for evidence-based rollout strategies that minimize drug resistance.

**Methods:** We used a stochastic compartmental model of men who have sex with men (MSM) in the United States (U.S.) to compare two main strategies—equal allocation and sequential drug deployment—for two new and one existing drug and measured the time for each drug to reach a resistance prevalence threshold of 5%. We conducted broad analyses assessing the sensitivity of our results to wide variation in parameters governing the baseline behavior of the model and drug resistance evolution and fitness.

**Results:** Compared to the equal allocation strategy, the sequential strategy had reached the resistance prevalence threshold i) for each drug individually in at least as many simulations; ii) for all three drugs in at least as many simulations; and iii) for at least as many drugs on average. Aber 10 years, no equal allocation strategy simulations had met the 5% resistance prevalence threshold for any of the drugs, whereas 99.6% of sequential simulations had for the first drug, of which 3.5% had also met the threshold for the second drug. The sequential strategy was worse for nearly every reasonable combination of model parameters.

**Conclusion:** In a model of U.S. MSM, the equal allocation strategy for introducing new drugs for gonorrhea matched or outperformed the strategy of sequential introduction in terms of resistance prevalence.

**Summary:** Immediately introducing two new antibiotics to treat gonorrhea alongside one currently used therapy is more effective at minimizing drug resistance than holding the new drugs in reserve.

## INTRODUCTION

The burden of disease caused by *Neisseria gonorrhoeae* and the emergence of extremely drug-resistant strains highlights the need for evidence to optimize treatment options. *N. gonorrhoeae* is an obligate human bacterial pathogen that causes the sexually transmitted infection (STI) gonorrhea. There were an estimated 82.4 million gonorrhea infections worldwide in 2020 and over half a million in the United States (U.S.) in 2023.^1,2^ High-risk demographics in the U.S. include young adults (especially ages 20-24), men who have sex with men (MSM), and those identifying as American Indian, Alaskan Native, or Black or African American.^2^ Gonorrhea treatment guidelines in the U.S. recommend a 500 mg intramuscular injection with cebriaxone, but strains resistant to cebriaxone have spread globally.^3–8^ Unlike other common STI pathogens, *N. gonorrhoeae* has evolved resistance to every antibiotic approved to treat it.^9^

Two new first-in-class antibiotics have recently reported positive phase III trial results for gonorrhea treatment: zoliflodacin and gepotidacin.^10,11^ Zoliflodacin is a single-dose oral regimen, while the gepotidacin regimen in the phase III study was two oral doses taken 10-12 hours apart. Upon their approval for gonorrhea treatment, recommendations will be required to guide clinical practice.

Modeling studies have examined similar questions about how best to roll out interventions for gonorrhea.^12–14^ Most relatedly, one study assessed multiple strategies for introducing one new antibiotic for gonorrhea using a deterministic modeling framework.^12^ It found that keeping the new drug in reserve until the currently used therapy reached its resistance prevalence threshold was consistently worse than strategies that deployed the new drug upon approval. Other studies have assessed interventions including point-of-care testing, rapid susceptibility testing, ^15–18^ vaccination,^13,14,19^ screening, and post-exposure prophylaxis.^20–22^ No study so far has considered the availability of two new drug options for the treatment of gonorrhea and used a stochastic framework aimed at capturing stochastic resistance emergence and extinction dynamics.

In this study, we compared rollout strategies for two new treatment options in addition to one existing therapy using a stochastic compartmental model of gonorrhea in U.S. MSM. We assessed which rollout strategy maximized the useful lifespan of the three potential treatment options and which combinations of parameters most influenced this result.

## METHODS

### Model Structure and Implementation

We used a compartmental model of gonorrhea transmission among U.S. MSM adapted from previous studies.^12,15,16,20^ Briefly, the model follows susceptible-infected-susceptible (SIS) dynamics. Treatment is successful unless there is resistance to the drug used, in which case treatment fails. We modeled three general drugs, A, B, and C, to represent cebriaxone, zoliflodacin, and gepotidacin, respectively, acknowledging that the precise parameters (e.g., likelihood of emergence of resistance and fitness cost) for these specific drugs remain unknown.

The population is stratified into high-, medium-, and low-risk, which describe the number of partnerships formed by individuals in each stratum. Mixing is not fully assortative; partnerships can be formed within one’s own risk stratum and outside of it. All compartments and transitions between them are shown in **Supplementary Figure 1** and described in **Appendix I & II**.

There are two sets of infected compartments: symptomatic (Y) and asymptomatic (Z). Symptomatic individuals are assumed to seek treatment, whereas asymptomatic individuals are only detected and treated at the rate of asymptomatic screening. There is an additional exit rate from symptomatic compartments representing the possibility of seeking care for retreatment if symptoms do not resolve aber failing treatment. Resistance can occur through two mechanisms in the model: 1) starting resistance is transmitted, meaning that previously uninfected individuals are directly infected with resistant bacteria, or 2) people infected with strains that are susceptible to the antibiotic with which they are treated develop resistance to that antibiotic. Resistance can only arise to one antibiotic at a time, as infected individuals are treated with a single antibiotic, though multi-drug resistance can emerge on already resistant backgrounds.

Transmission, care seeking, screening, natural recovery, and resistance emergence were modeled as a continuous time Markov chain with event rates (**Appendix II**) matching those of the deterministic system (**Appendix I**). The system was then simulated in discrete time using binomial τ-leaping with a time step of one day.^23,24^ The implementation of this approach involved multinomial draws from each compartment at each time step. This stochastic framework captures rare resistance emergence events and lineage extinctions, which govern differences in dynamics between different rollout strategies.^25^

The models were implemented in R 4.4.1 using the Odin package^26^ and run on the FASRC Cannon high performance computing cluster at Harvard University. All code used in this project can be found at: https://github.com/gradlab/MK_twodrugintro_public.

### Strategy Comparison

We compared two drug rollout strategies: sequential and equal allocation. In each strategy, individuals are treated with one drug at a time. At the beginning of the sequential strategy, everyone is treated with drug A. Once the resistance prevalence threshold is met for drug A, it phases out and is replaced by B. Once the resistance prevalence threshold is met for drug B, it phases out and is replaced by C until the end of the simulation. At the beginning of the equal allocation strategy, each infected individual has a 1/3 chance of being treated with drug A, drug B, or drug C. Once the first drug hits its resistance prevalence threshold, it is no longer used; treatment is then either of the two remaining drugs, each with a 1/2 chance. When the resistance prevalence threshold is met for the second drug, it phases out. The third drug is then used for the rest of the simulation (**Supplementary Figure 2**).

In both strategies, people with symptomatic infections have a chance to seek retreatment, in which case they are treated with a last resort treatment that is not drugs A, B, or C, such as ertapenem.^5^ Retreatment is assumed to always be successful. We did not explore combination therapies because the interactions among the drugs have not been studied in clinical senngs and approval for combination therapy is not being pursued.

The resistance prevalence threshold was set to 5% in the baseline scenario, in keeping with longstanding guidelines.^27,28^ However, we also explored scenarios where the threshold was 1% or 10%.

### Parameter Selection and Sensitivity Analysis

Model parameters were selected by literature search, maximum likelihood estimation calibration, and sensitivity analysis (**Supplementary Table 1, Supplementary Table 2**). Parameters were divided into two categories: those governing underlying transmission dynamics (Group 1), and those relating to drug resistance emergence and persistence (Group 2). For Group 1 parameters, which were unlikely to affect strategy ordering, we identified starting values from the literature and calibrated a drug A-only model run for two years using the R bbmle package as described.^12,29^ We then conducted a Latin hypercube sampling (LHS) sensitivity analysis over all possible values of the calibrated parameters to increase our confidence in the calibrated parameter values.

We did not calibrate Group 2 parameters and instead performed a broad LHS sensitivity analysis (**Supplementary Table 2**). These parameters included the probability of resistance emerging on treatment, the relative transmissibility cost (relative fitness) of drug resistance, and the starting number of cases of resistance in the system. While these quantities are difficult to ascertain empirically, we can roughly bound the probability of resistance emerging on treatment using the substitution rate and observed treatment failures. The *N. gonorrhoeae* substitution rate is on the order of 10^−6^ substitutions per site per year in magnitude.^30^ This is the rate of neutral mutations that have established, and therefore a lower bound on the order of magnitude for our model’s resistance emergence probability parameter as mutations that are selected are more likely to establish. While the upper bound is more challenging, a resistance emergence probability of 10^−4^ or above would mean that there were more than one treatment failures per 10000 cases, which would amount to at least 60 treatment failures in the US in 2023. To be conservative, we explored mutation probabilities one order of magnitude outside of what we thought most likely (10^−3^ - 10^−7^).

We used LHS (the lhs package in R^31,32^) for both sensitivity analyses to effciently and comprehensively survey a multidimensional parameter space. For each sensitivity analysis, we generated 1000 unique sets of parameter combinations. For each parameter combination, we ran 100 stochastic simulations of each strategy.

## RESULTS

### Baseline Scenario

Models run with the baseline parameter set showed that the equal allocation strategy was consistently better than the sequential strategy (**Figure 1**). At each time step, the same number or fewer simulations had met the 5% resistance prevalence threshold to each of the three drugs under the equal allocation strategy compared to the sequential strategy. By year five, no equal allocation strategy simulations had met the threshold for any of the drugs, but 61.5% of sequential strategy simulations had reached 5% resistance prevalence for drug A. By year 10, no simulations for the equal allocation strategy had met the threshold for any of the drugs, but 99.6% of sequential strategy simulations had reached 5% resistance prevalence for drug A, and 3.5% of sequential strategy simulations had reached 5% resistance prevalence for drug B. Aber 30 years, 25.2% of equal allocation strategy whereas 100% of sequential strategy simulations had reached 5% resistance prevalence for drug A, 75.1% of sequential strategy simulations had reached 5% resistance prevalence for drug

**Figure 1:**
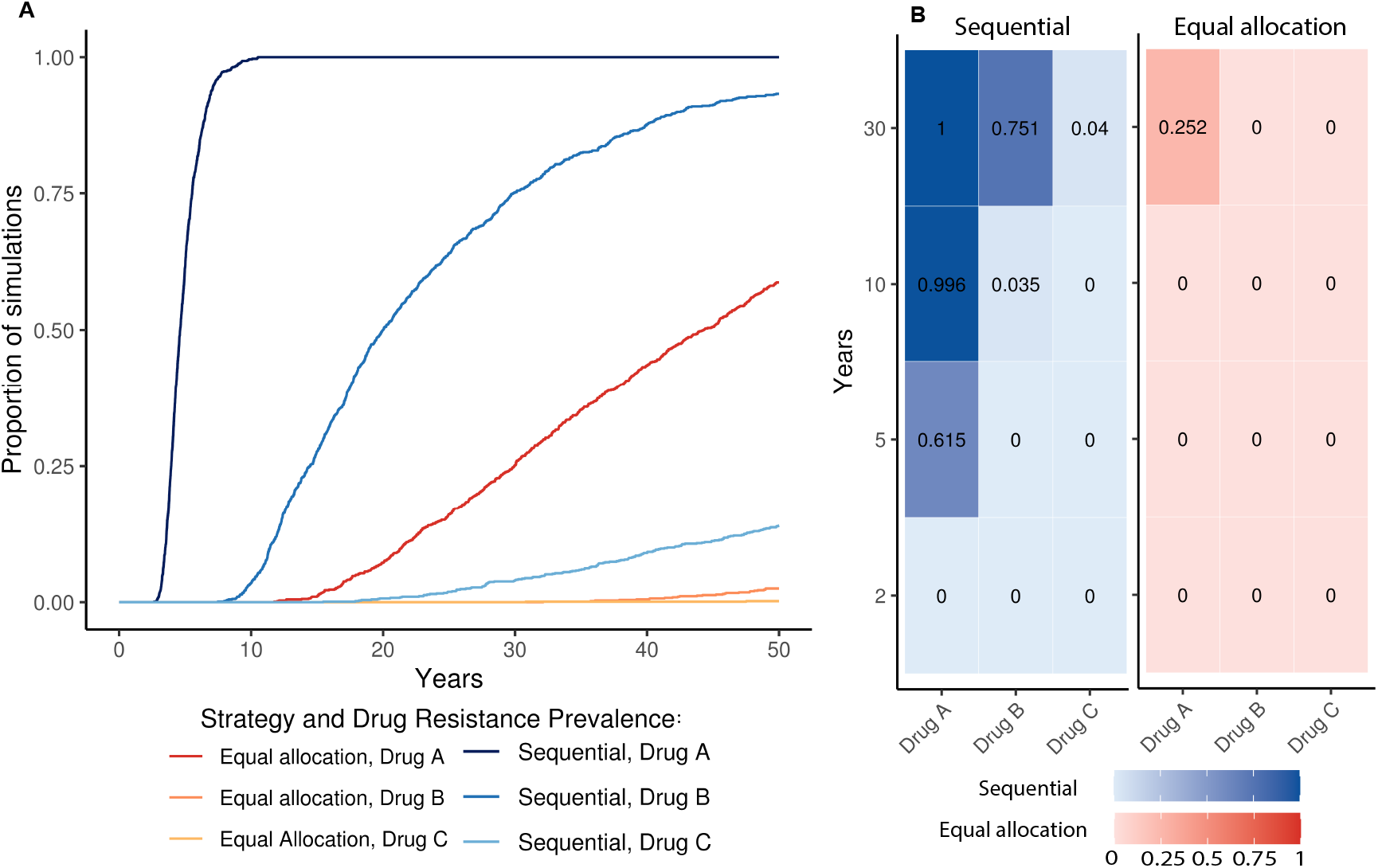
Proportion of stochastic simulations reaching the 5% prevalence threshold over time under the equal allocation and sequential strategies. Results from 1000 simulations of each strategy using baseline parameters. A: The proportion of the 1000 simulations of each strategy that have reached the 5% prevalence threshold at each time point. Blues colors show the trajectories for drugs A, B, and C in the sequential strategy, and red colors show the trajectories for drugs A, B, and C in the equal allocation strategy. B: The proportion of the 1000 sequential strategy simulations (blue, leb) and the 1000 equal allocation strategy simulations (red, right) that have met the 5% resistance prevalence thresholds for drugs A, B, and C (X-axis) aber 2, 5, 10, and 30 years of simulation (Y-axis).

At every time point, the sequential strategy had lost at least as many of the three available drugs on average as the equal allocation strategy. At two, five, and 10 years, none of the equal allocation strategy simulations had lost any drugs whereas by year five, sequential strategy simulations had lost 0.62 drugs on average (38.5% of simulations lost no drugs, 61.5% of simulations had lost 1 drug, **Supplementary Table 3**) and by year 10, sequential strategy simulations had lost an average of 1.03 drugs (0.4% of simulations had lost no drugs, 96.1% of simulations had lost 1 drug, 3.5% lost 2). By year 30, the equal allocation strategy had lost an average of 0.25 drugs (74.8% had lost no drugs, 25.2% had lost 1 drug), and the sequential strategy had lost on average 1.83 (24.9% had lost 1 drug, 71.1% had lost 2 drugs, 4% had lost all 3 drugs, **Supplementary Figure 3**).

Similarly, across the study duration, the sequential strategy had lost all three drugs in the same or a greater proportion of simulations compared to the equal allocation strategy. On average, a small proportion of simulations from either strategy had lost all three drugs, but aber 50 years, no equal allocation strategy runs had lost all three drugs compared to 14.0% of sequential strategy runs (**Supplementary Figure 4**).

The equal allocation strategy also outperformed the sequential strategy at resistance prevalence thresholds of 1% and 10%, with an equal or greater average number of drugs lost at each time point in the sequential strategy (**Figure 2**).

**Figure 2:**
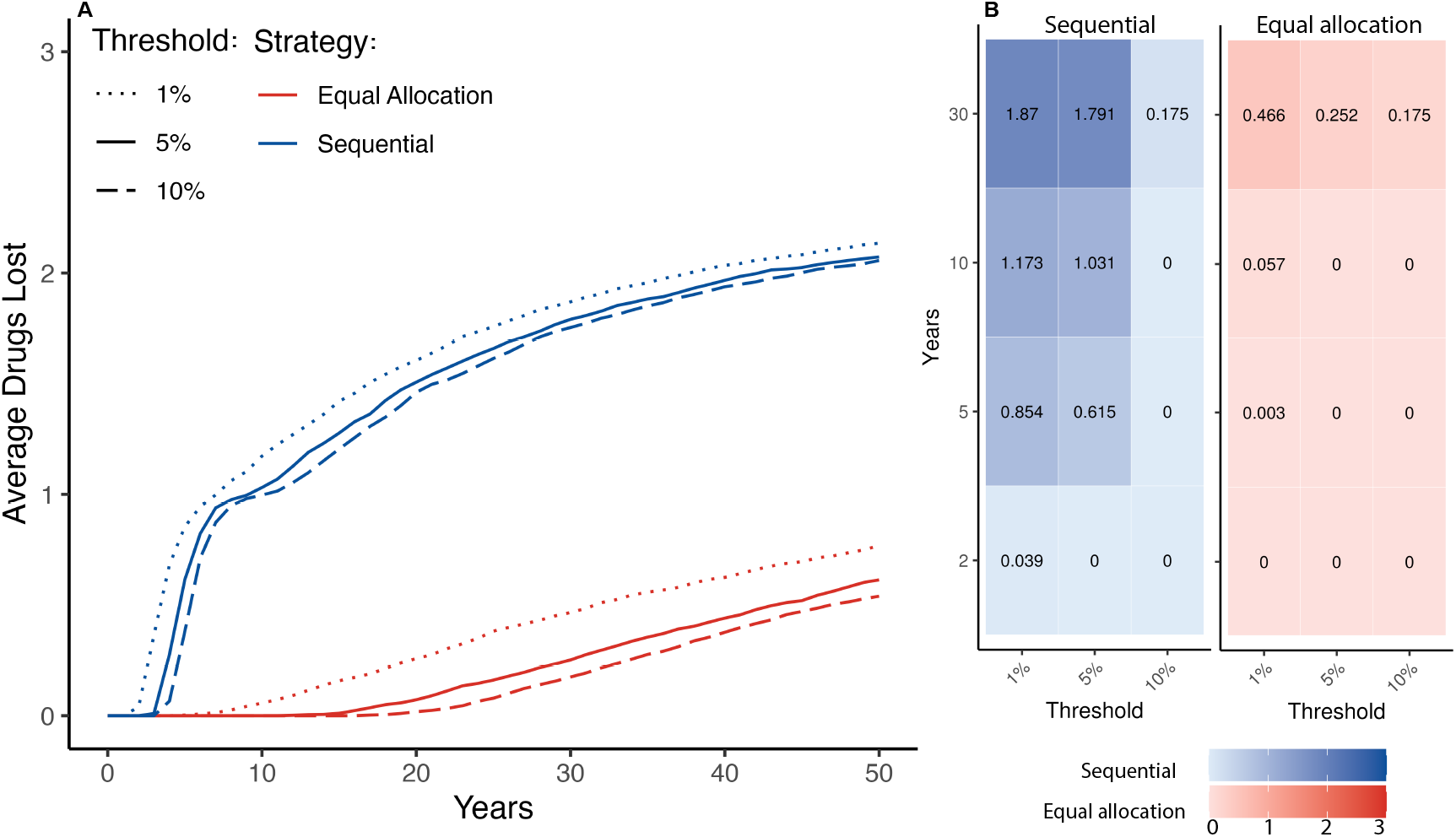
Average drugs lost over time across stochastic simulations by strategy and threshold. The threshold refers to the prevalence of resistance required to prompt a change in treatment. In the sequential strategy, once the threshold is met, the drug is no longer used, and a new drug is phased in. In the equal allocation strategy, the drug is no longer used and the probability of using the remaining drug or drugs increases. A: Average number of drugs lost across 1000 stochastic simulations of the sequential and equal allocation strategies. Blue lines indicate the average number of drugs lost across the 1000 sequential strategy simulations, and red lines indicate the average number of drugs lost across the 1000 equal allocation strategy simulations. Dotted lines indicate that the threshold for that set of simulations across both strategies was 1%, solid lines indicate a 5% threshold, and dashed lines indicate a 10% threshold. B: The average number of drugs lost in the sequential strategy (blue, leb) and the equal allocation strategy (red, right) aber 2, 5, 10, and 30 years of simulation.

### Sensitivity analyses

In each of the 1000 Group 1 parameter combinations (**Supplementary Table 1**), the sequential strategy had lost each drug in an equal or greater proportion of simulations than the equal allocation strategy (**Supplementary Figure 5**). The results from the sensitivity analysis varying Group 2 parameters (**Supplementary Table 2**) identified only a small number of scenarios where more drugs were lost on average in the equal allocation strategy compared to the sequential strategy (**Figure 3**). By year 10, simulations from only 22 parameter combinations (2.2% of all combinations) showed on average more drugs lost under the equal allocation strategy. In each of these parameter combinations, none of the sequential strategy simulations lost any drugs, and a maximum of 6% of equal allocation strategy simulations had lost either drug B or drug C (**Figure 3, Supplementary Table 4, Supplementary Figures 6 & 7**).

**Figure 3:**
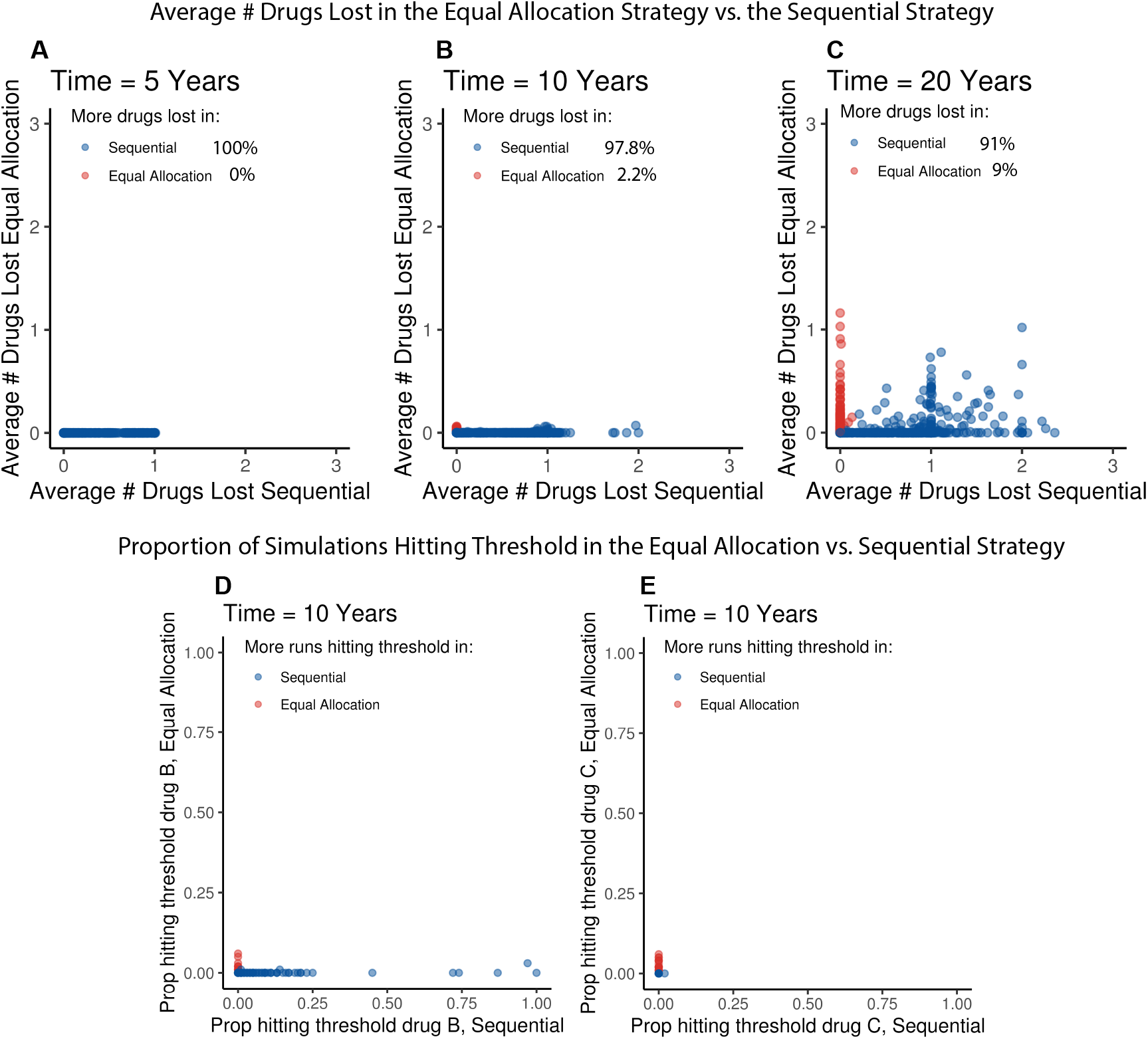
Results from sensitivity analysis varying parameters that may influence strategy ordering. Each dot represents one of the 1000 Latin hypercube sampled parameter combinations with 100 simulations run under the sequential strategy and 100 simulations run under the equal allocation strategy. Panels A-C: The X-axis represents the average number of drugs lost across the 100 sequential simulations with that set of parameters, and the Y-axis represents the average number of drugs lost across the 100 equal allocation simulations with that set of parameters at 5 (A), 10 (B), and 20 (C) years. Dots are colored blue if more drugs were lost in the sequential strategy, and red if more drugs were lost in the equal allocation strategy. Insets indicate the percentage of 1000 parameter combinations where more drugs were lost in the equal allocation strategy on average than in the sequential strategy at that time point. Panels D & E: The X-axis represents the proportion of the 100 sequential strategy simulations that had met the resistance prevalence threshold to drug B (panel D) or drug C (panel E), and the Y-axis represents the proportion of the 100 equal allocation strategy simulations that had met the resistance prevalence threshold to drug B (panel D) or drug C (panel E) aber 10 years.

Aber filtering out parameter combinations that included probabilities of resistance emergence greater than or equal to 10^−4^, there were 7 parameter combinations where more drugs had been lost on average under equal allocation than under sequential aber 10 years. In each of these scenarios, only 1-2% of equal allocation simulations had reached 5% resistance prevalence for either drug B, or drug C, while none of the drug resistance prevalence thresholds were met in the sequential simulations (**Supplementary Figure 7**). Thus, over nearly all parameter space, the sequential strategy lost more drugs on average than the equal allocation strategy.

## DISCUSSION

In our modeling study, the equal allocation strategy, in which the two new antibiotics and the currently used therapy are deployed in equal proportion, randomly across the population treated, minimized the emergence and spread of antibiotic resistance when compared to a strategy of rolling out the new drugs sequentially. Under the baseline set of parameters, resistance to all three treatments was less likely to reach a 5% prevalence threshold in the equal allocation strategy than in the sequential strategy. This result held at resistance prevalence thresholds for drug loss of 1% and 10%. Across the simulation period, more drugs on average were lost due to antimicrobial resistance under the sequential strategy than under the equal allocation strategy. Our results were robust to a wide range of parameters that alter gonorrhea transmission dynamics.

The model results were most sensitive to the number of cases of starting resistance, the probability of resistance emergence, and the relative fitness. Our sensitivity analysis showed that it was possible to derive parameter combinations where the equal allocation strategy performed worse than the sequential strategy, but this required extreme and unlikely combinations of parameter values. Of the 22 parameter combinations where the equal allocation strategy had lost, on average, more drugs than the sequential strategy by year 10, only a small proportion of simulations had lost any drugs under equal allocation; a maximum 6% of simulations had met the 5% prevalence threshold for drug B or drug C. In these scenarios, drug A never reached its resistance prevalence threshold and therefore drugs B and C were never used in the sequential strategy; in contrast, their use in the equal allocation strategy meant that in some scenarios emerging or preexisting resistance to these drugs propagated and reached the threshold.

We classified the majority of the 22 parameter combinations where equal allocation was worse into “Class I”, where at least one emergence probability was greater than 10^−4^ (**Supplementary Figure 6**). For example, in one parameter combination, the probability of resistance emerging on treatment with drug B was 2.86 × 10^−4^, which would mean that, for 600,000 cases of gonorrhea treated with drug B, about 200 would fail treatment, which is an improbably high treatment failure rate that would likely draw attention. The seven remaining parameter combinations where the resistance emergence probability for all drugs was less than 10^−4^ (Class II) required: i) a relatively high number of starting cases of B or C resistance that propagated in the equal allocation strategy but could not persist in the sequential strategy, ii) a high relative fitness of B or C resistance (close to 1), and iii) a low enough relative fitness and or resistance probability for drug A such that the resistance prevalence threshold for drug A was never met and therefore drugs B and C were never used under the sequential strategy (**Supplementary Table 4, Supplementary Figure 7**). Even when this occurred, only 1-2% of simulations had reached the resistance prevalence threshold for drugs B or C aber 10 years under the equal allocation strategy. This emphasizes that the cases where the sequential strategy outperformed the equal allocation strategy were edge cases involving a co-occurrence of unlikely events.

Several factors not considered in this study may impact rollout decisions. Here, we focused on an MSM-only sexual network in the U.S., thus these results may not generalize to other sexual network structures or country senngs with higher baseline cebriaxone resistance prevalences and different asymptomatic screening practices. Considerations of drug price, availability, preferences for modes of administration, and toxicities were not included but will be important drivers of drug use and resistance patterns that will also depend on context.

We modeled a closed system with no importation of drug resistance and did not include the effects of bystander selection for drug resistance due to antibiotic use for indications outside of gonorrhea. We assumed that all drug resistance caused treatment failure and did not model within-individual evolution and variation that may occur along a transmission chain. We fixed parameter values in each simulation, so that, for example, the relative fitness of a particular pattern of resistance did not change over time. Our probability of resistance emergence on treatment broadly referred to any kind of genetic event leading to resistance but did not differentiate between point mutations or recombination events, which could vary by drug and have different rates. There is also substantial uncertainty surrounding the parameter values governing our models. However, we note that our results held across nearly all possible combinations of the parameter values and form a baseline intuition about drug resistance dynamics to be expanded upon in the future.

Our framework can be used for further investigations, including evaluating strategies where all drugs are available, but some are used more than others, the impact of drug synergy and cross resistance, given that zoliflodacin and gepotidacin target the same molecular complex as fluoroquinolones, and adaptive strategies that change in response to data on the likelihood of emergence of resistance and the relative fitness cost of resistance. Alternative screening strategies could also meaningfully impact resistance minimization; future studies building on our work could explore expedited partner therapy, rapid gonorrhea tests and rapid susceptibility testing, and vaccine usage.

In conclusion, introducing two new antimicrobials to treat gonorrhea alongside the currently used treatment was more effective at minimizing drug resistance than introducing the drugs in sequence. Our results reinforce that distributing the selective pressures across the population minimizes the emergence of drug resistance (**Supplementary Figure 8**). This aligns with deterministic modeling evaluating the introduction of one new treatment for gonorrhea, and malaria modeling showing that using multiple first-line therapies is beneficial.^12,33^ While traditional antimicrobial stewardship emphasizes reserving antibiotics, these results illustrate the importance of disease and context-specific decision-making.

## Data Availability

All data produced in the present work are contained in the manuscript

https://github.com/gradlab/MK_twodrugintro_public

## Author Contribu1ons

MCK designed and implemented the study, analyzed the results, and drabed the manuscript. KOR contributed to the design and implementation, supervised the analysis, and edited the manuscript. DH contributed to the implementation and analysis and edited the manuscript. ER contributed to the implementation and edited the manuscript. YHG contributed to the design, conceptualization, and analysis of results, edited the manuscript, and supervised the project. The authors thank all members of the Grad Lab for their valuable feedback on the project, especially QinQin Yu, and thank Jeff Imai-Eaton and Marc Lipsitch for their input.

