## Supplemental Materials for "Comparing Strategies to Introduce Two New Antibiotics for Gonorrhea: A Modeling Study"

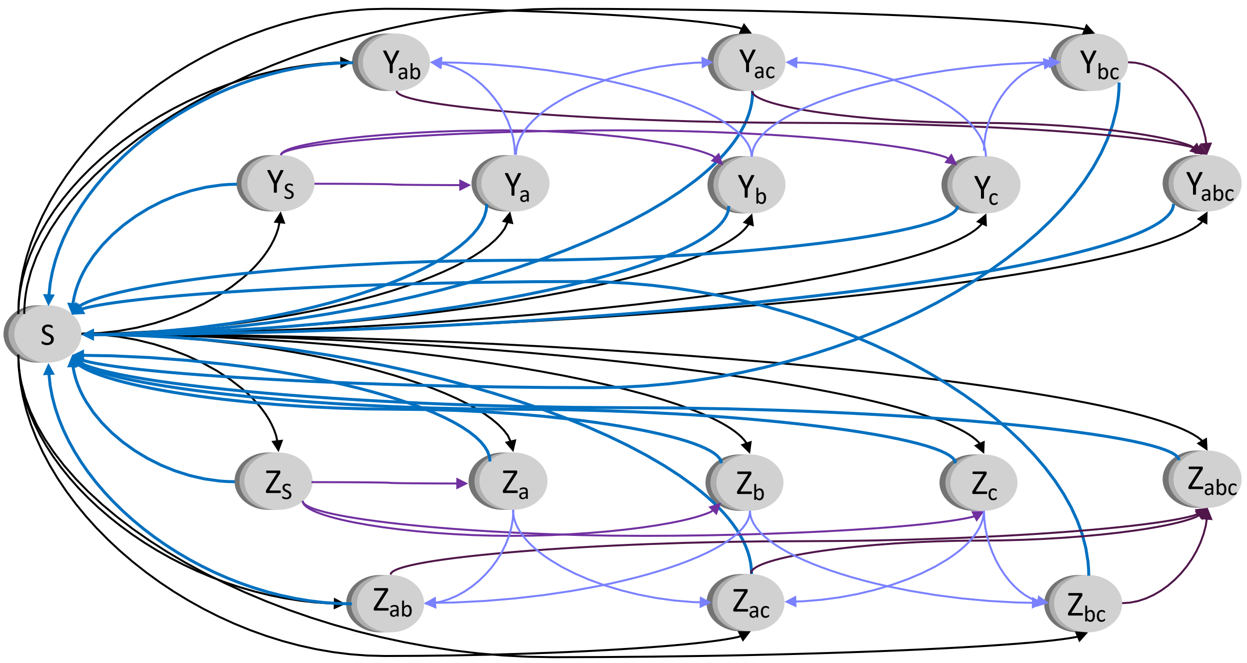


**Supplementary Figure 1: Model schematic**. Grey circles represent model compartments and arrows show possible flows in and out of compartments. Each circle has three layers indicating the three risk strata. Black arrows show flows from the uninfected, susceptible compartment (S) to infected compartments either symptomatic (Y), or asymptomatic (Z). Light blue arrows show recovery from infected compartments (Y or Z) back to being uninfected and susceptible (S). Dark purple arrows show flow from drug sensitive compartments ($Y_s$ and $Z_s$) to single resistance compartment ($Y_a$, $Y_b$, $Y_c$, and $Z_a$, $Z_b$, $Z_c$). Light purple arrows show evolution of double resistance ($Y_{ab}$, $Y_{ac}$, $Y_{bc}$, and $Z_{ab}$, $Z_{ac}$, $Z_{bc}$) upon a single resistance background, and dark blue arrows show triple resistance evolution ($Y_{abc}$, and $Z_{abc}$).

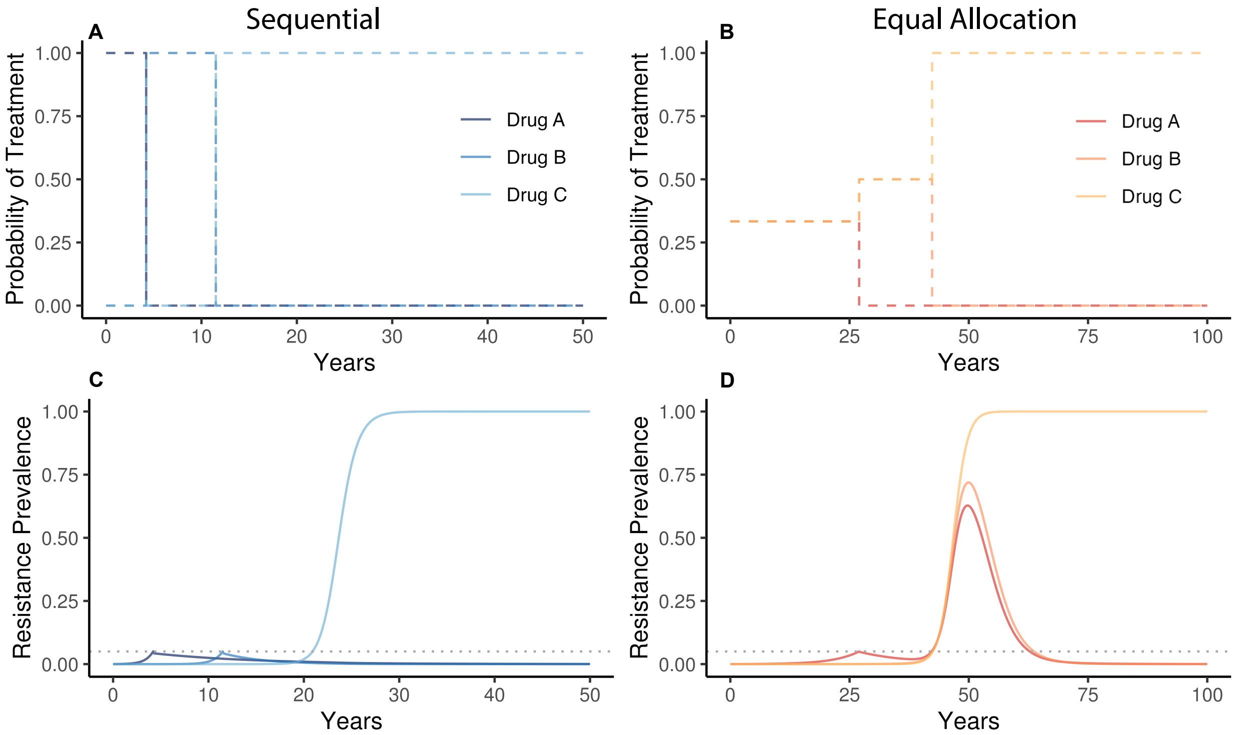


**Supplementary Figure 2**: **Treatment probabilities in sequential and equal allocation strategies.** Deterministic model output using baseline parameters to illustrate that the probability of treatment with each drug depends on the prevalence of resistance to that drug in the system. We say that a drug has been lost once it has reached the resistance prevalence threshold. A. Probability of treatment with drugs A, B, and C over the course of a 50-year simulation using the sequential strategy. B. Probability of treatment with drugs A, B, and C over the course of a 100-year simulation using the equal allocation strategy. 100 years are shown for the equal allocation strategy for demonstrative purposes to observe the model as it equilibrates. C. Prevalence of resistance to drugs A, B, and C over the course of the sequential simulation. D. Prevalence of resistance to drugs A, B, and C over the course of the equal allocation simulation.

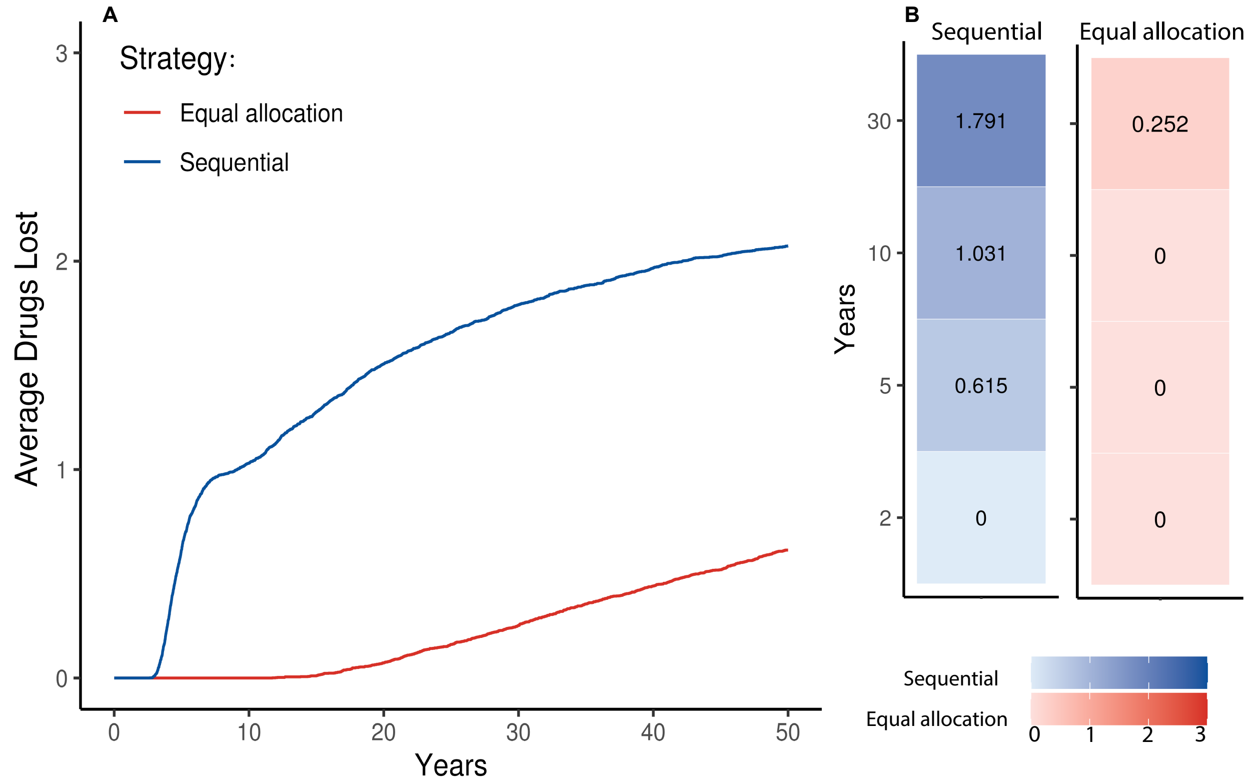


**Supplementary Figure 3: Average number of drugs that have hit the 5% prevalence threshold by strategy.** Results are shown from 1000 stochastic simulation each of the equal allocation and sequential strategies. A: The average number of drugs that have reached the 5% resistance prevalence across the 1000 simulations for the sequential strategy (blue) and the equal allocation strategy (red). B: The average number of drugs that have reached the 5% resistance prevalence threshold for the sequential strategy (blue, left) and the equal allocation strategy (red, right) at 2, 5, 10, and 30 years.

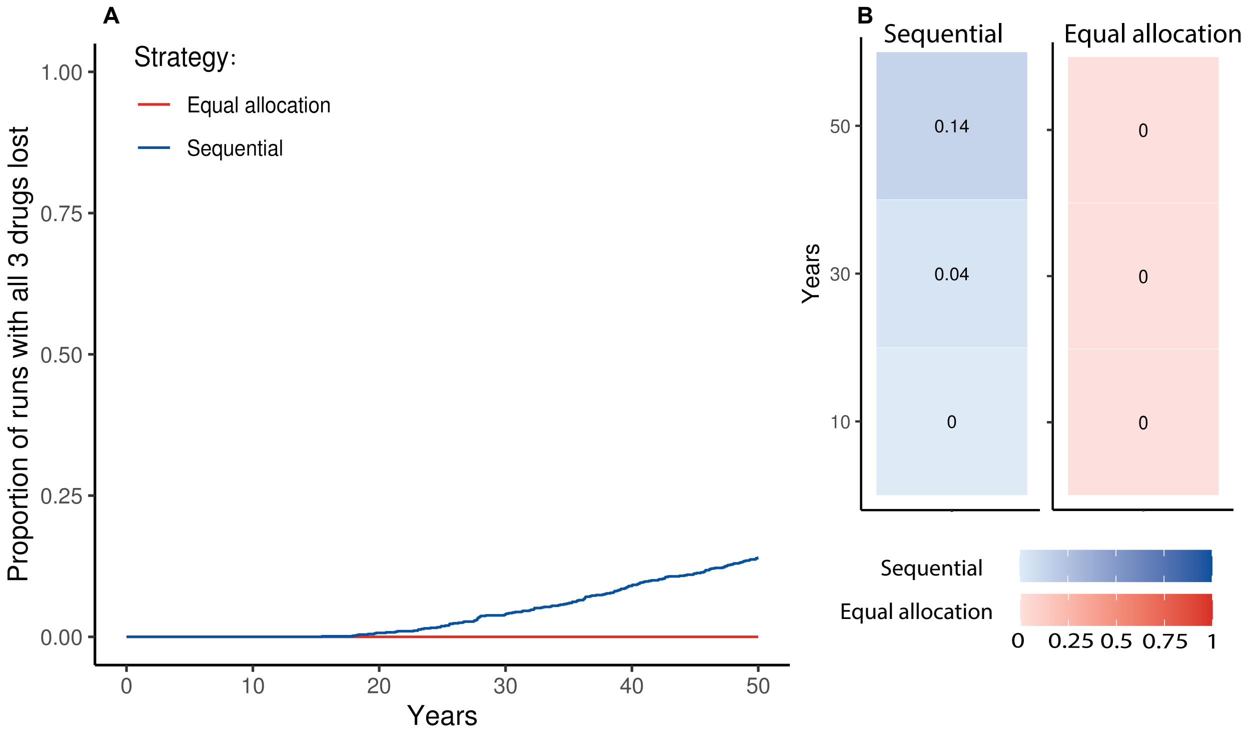


**Supplementary Figure 4: Proportion of stochastic simulations reaching the 5% prevalence threshold for all three drugs over time under both strategies.** Results are shown from 1000 stochastic simulation each for the equal allocation and sequential strategies. A: The proportion of simulations where resistance to drugs A, B, and C has reached the 5% prevalence threshold for the sequential strategy (blue) and the equal allocation strategy (red). B: The proportion of the 1000 sequential strategy simulations (blue, left) and the 1000 equal allocation strategy simulations (red, right) that have met the 5% resistance prevalence thresholds for all drugs A, B, and C (X-axis) after 10, 30 and 50 years.

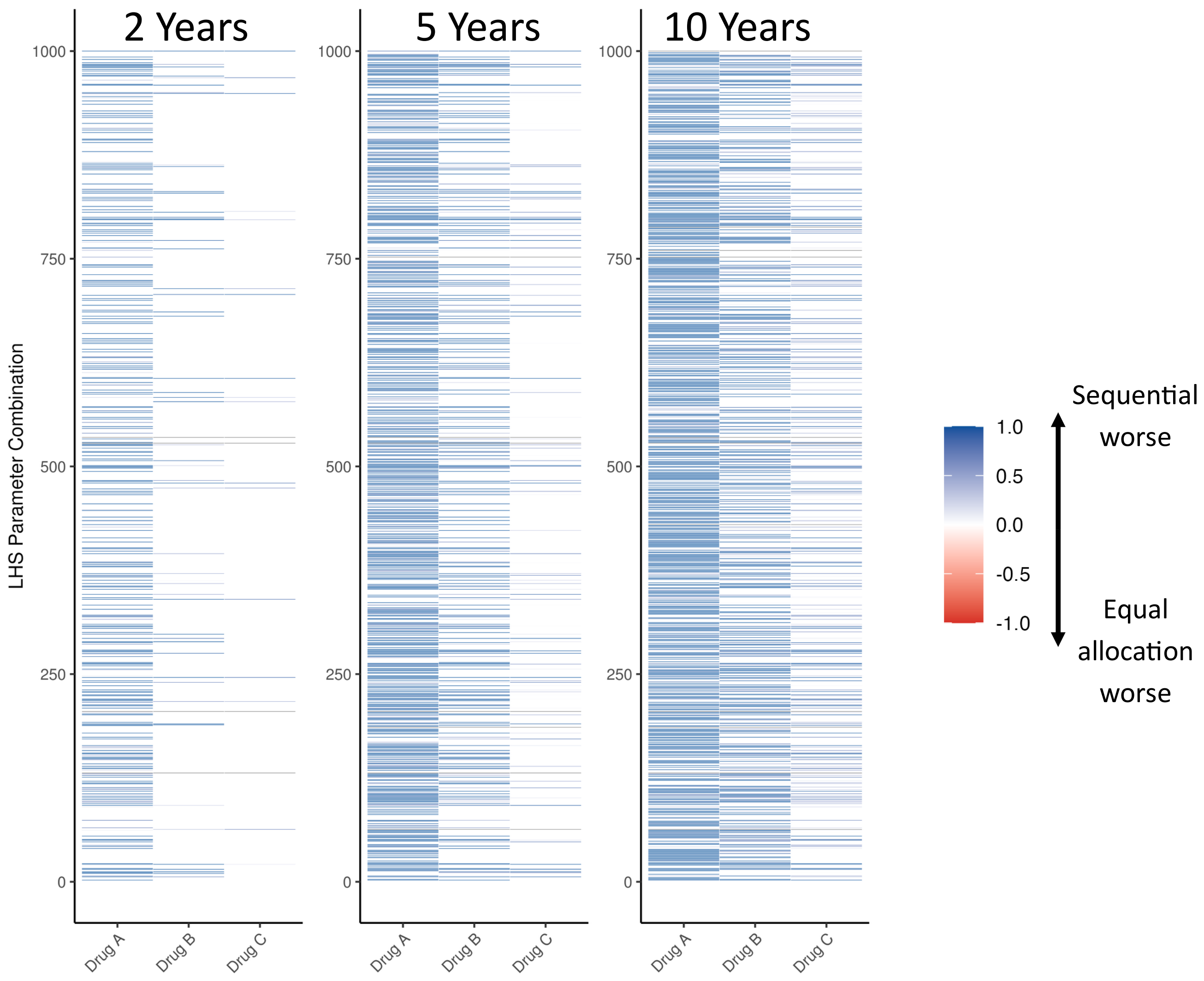


**Supplementary Figure 5: Difference in proportion of simulations meeting resistance prevalence threshold for each drug between strategies varying Class I parameters**. Y-axis indicates which of the 1000 Latin hypercube sampling (LHS) parameter combinations varying Class I parameters was run for 100 simulations under the equal allocation and 100 simulations under the sequential strategy. Blue indicates that a greater proportion of sequential strategy simulations have met the 5% resistance prevalence threshold for a given drug compared to equal allocation, red indicates that a greater proportion of simulations have met resistance prevalence threshold in equal allocation than sequential.

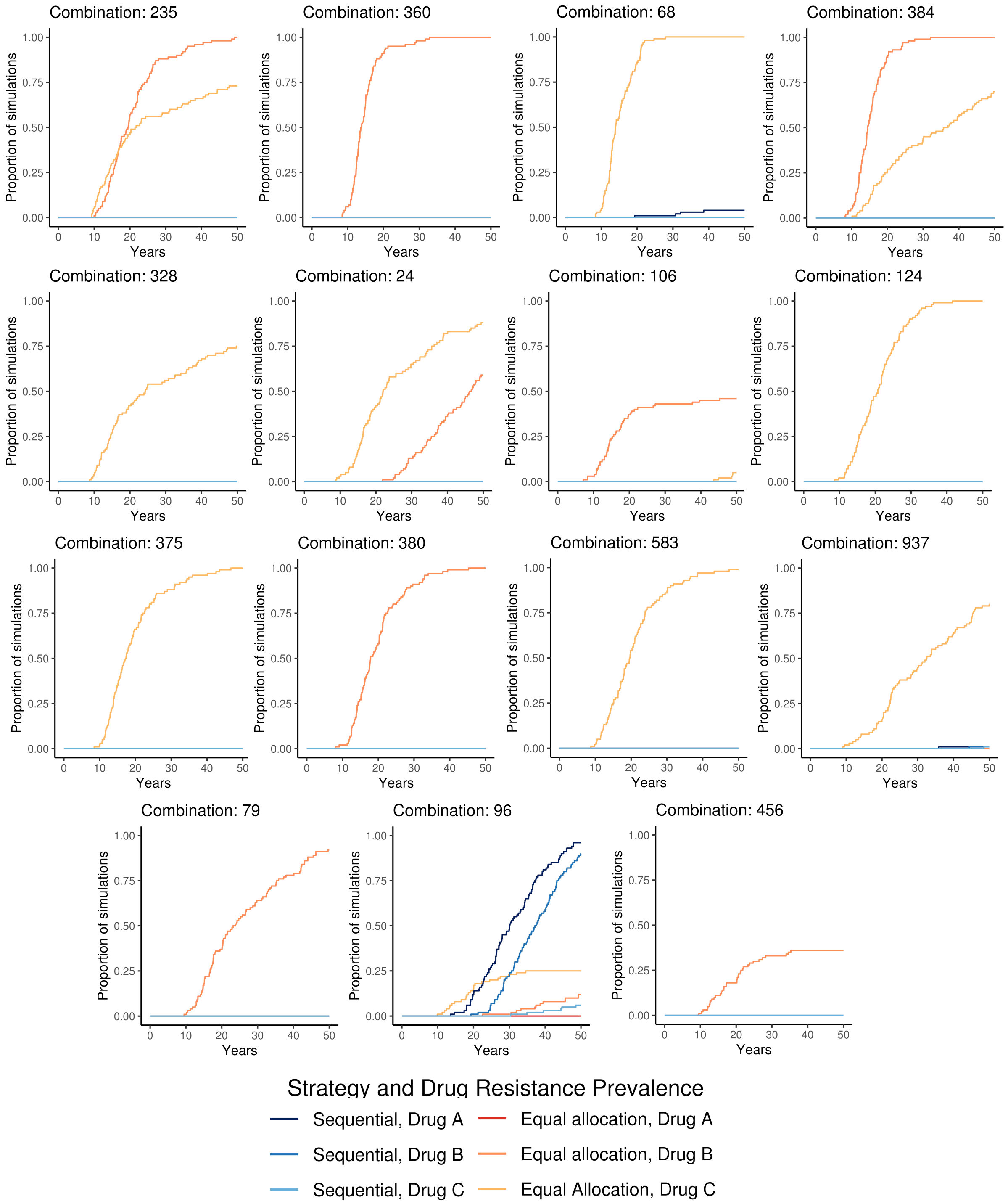


**Supplementary Figure 6: Class I parameter combinations from Latin hypercube sensitivity analysis**. 15 parameter combinations from Latin hypercube sampling (LHS) with more drugs lost an average in the equal allocation strategy than in the sequential strategy after 10 years. Parameter values can be found in Supplementary Table 4. For each LHS parameter combination, the proportion of simulations hitting resistance prevalence threshold for each drug and strategy is plotted on the same set of axes.

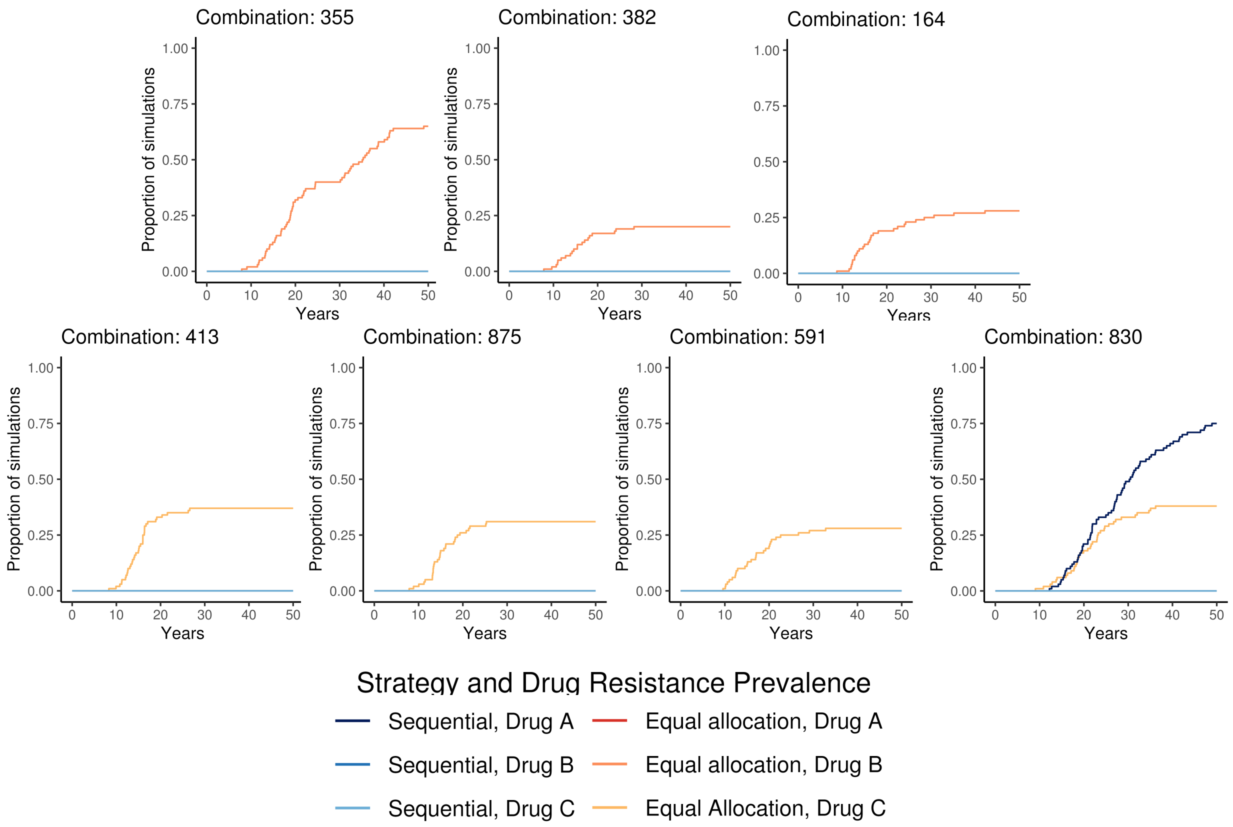


**Supplementary Figure 7: Class II parameter combinations from Latin hypercube sampling sensitivity analysis**. Seven Latin hypercube sampling (LHS) parameter combinations where equal allocation was worse than sequential after filtering mutation probabilities greater than $10^{-4}$. Parameter values can be found in Supplementary Table 4. For each LHS parameter combination, the proportion of simulations hitting resistance prevalence threshold for each drug and strategy is plotted on the same set of axes.

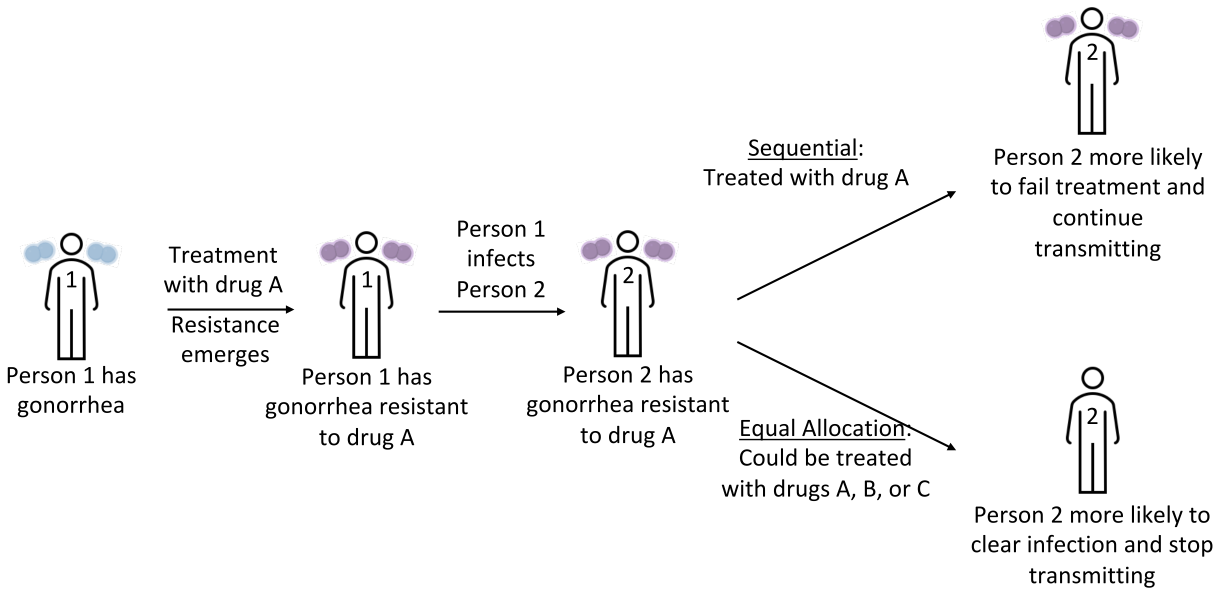


**Supplementary Figure 8: Framework schematic**. The equal allocation strategy more effectively distributes selective pressures across the population, making it harder for *N. gonorrhoeae* resistance to survive and propagate.

**Supplementary Table 1: Group 1 Model Parameter Values**: Parameters governing model structure, sexual partnership formation and behavior, and infection characteristics.

| Parameter | Description | Baseline value | Range explored | Calibration starting value | Source |
|---|---|---|---|---|---|
| Gonorrhea starting prevalence | Calibration target | 3% | -- | | [12,20,34] |
| N | Population size | $10^6$ | Fixed | | Assumption |
| $N_k$ | Size of sexual activity risk groups | | Fixed | | [12,15,20] |
| $N_1$ | Low risk | $3 \times 10^5$ | | | |
| $N_2$ | Intermediate risk | $6 \times 10^5$ | | | |
| $N_3$ | High risk | $1 \times 10^5$ | | | |
| $\theta$ | Partner change rate (per year) – lowest risk group | 1.38 | 0.2 – 365 | 1.22 | Model fitting, [12,20] |
| | Partner change rate (per year) – intermediate risk group | $5*\theta$ | | | |
| | Partner change rate (per year) – high risk group | $20*\theta$ | | | |
| $\varepsilon$ | Mixing parameter | 0.24 | 0 – 1 | 0.22 | Model fitting, [12,20] |
| $\sigma$ | Proportion of incident infections with symptoms | 0.49 | 0 – 1 | 0.5 | Model fitting, [20,35] |
| b | Probability of transmission per partnership | 0.52 | 0 – 1 | 0.5 | Model fitting, [12,20,21] |
| $\beta$ | Per capita transmission matrix | -- | -- | -- | As described in supplement of [12] |

| | | | | | |
|---|---|---|---|---|---|
| $1/\delta$ | Duration of recovery from untreated infection (natural recovery, days) | 91.84 | 1 – 365 | 105 | Model fitting, [20,36,37] |
| $\frac{1}{T_s}$ | Average time to test and treat, symptomatic infection (days) | 14.94 | 1 – 90 | 15 | Model fitting, [20] |
| $\frac{1}{T_{sr}}$ | Average time to retreatment, symptomatic infection (days) | $\frac{1}{T_s} * 3$ | $\frac{1}{T_s} * [1 - 10]$ | -- | [12,15,20] |
| $\frac{1}{T_m}$ | Average time to screen and treat, asymptomatic infection (days) | 879.47 | 365/(0 – 1) | 912.5 | Model fitting, [12,20,38] |
| $\kappa$ | Proportion retreated after initial treatment failure, symptomatic | 0.90 | 0 – 1 | -- | |
| $\xi_A, \xi_B, \xi_C$ | Probability of treatment (with drug A, B, or C) | Strategy dependent | -- | -- | -- |

**Supplementary Table 2: Group 2 Model Parameter Values**: Parameters that determine how likely mutations conferring resistance causing treatment failure are to arise, how fit drug resistant lineages are, and how much starting resistance was in the system at the beginning of the simulation.

| Parameter | Description | Baseline value | Range explored | Source |
|---|---|---|---|---|
| $\omega_A$ | Probability of emergence of resistance to A upon treatment with A | $10^{-4}$ | $10^{-3}$ - $10^{-7}$ | [30] |
| $\omega_B$ | Probability of emergence of resistance to B upon treatment with B | $10^{-5}$ | $10^{-3}$ - $10^{-7}$ | [30] |
| $\omega_C$ | Probability of emergence of resistance to C upon treatment with C | $10^{-6}$ | $10^{-3}$ - $10^{-7}$ | [30] |
| $f_A$ | Relative fitness (transmissibility) of single resistance to A compared to susceptible | 0.98 | 0.5 – 1 | [12,15] |
| $f_B$ | Relative fitness (transmissibility) of single resistance to B compared to susceptible | 0.95 | 0.5 – 1 | |
| $f_C$ | Relative fitness (transmissibility) of single resistance to C compared to susceptible | 0.95 | 0.5 – 1 | |
| $f_{AB}$ | Relative fitness (transmissibility) of AB dual resistance | $f_A * f_B$ | -- | Assumption |

| | | | | |
|---|---|---|---|---|
| | compared to susceptible | | | |
| $f_{AC}$ | Relative fitness (transmissibility) of AC dual resistance compared to susceptible | $f_A * f_C$ | -- | Assumption |
| $f_{BC}$ | Relative fitness (transmissibility) of BC dual resistance compared to susceptible | $f_B * f_C$ | -- | Assumption |
| $f_{ABC}$ | Relative fitness (transmissibility) of ABC triple resistance compared to susceptible | $f_A * f_B * f_C$ | -- | Assumption |
| resA | Starting cases of A resistance | 2 | 0 – 10 | [8] |
| resB | Starting cases of B resistance | 0 | 0 – 10 | |
| resC | Starting cases of C resistance | 0 | 0 – 10 | |

**Supplementary Table 3:** Number of simulations with baseline parameter set that had lost (reached the 5% resistance prevalence threshold for) 0, 1, 2, or 3 drugs at the specified time.

| Years | # Simulations with 0 drugs lost | # Simulations with 1 drug lost | # Simulations with 2 drugs lost | # Simulations with 3 drugs lost | Strategy |
|---|---|---|---|---|---|
| 2 | 1000 | 0 | 0 | 0 | Sequential |
| | 1000 | 0 | 0 | 0 | Equal Allocation |
| 5 | 385 | 615 | 0 | 0 | Sequential |
| | 1000 | 0 | 0 | 0 | Equal Allocation |
| 10 | 4 | 961 | 35 | 0 | Sequential |
| | 1000 | 0 | 0 | 0 | Equal Allocation |
| 30 | 0 | 249 | 711 | 40 | Sequential |
| | 748 | 252 | 0 | 0 | Equal Allocation |

**Supplementary Table 4: Parameter combinations where equal allocation was worse than sequential after 10 years.** Seven parameter combinations from Category 2 Latin hypercube sensitivity analysis where equal allocation strategy simulations had lost on average more drugs than sequential did after removing combinations with mutation probabilities greater than $10^{-4}$. “Diff” refers to the average number of drugs lost in the sequential strategy minus the average number of drugs lost in the equal allocation strategy.

| LHS combination | Diff | ResA | ResB | ResC | $\omega_A$ | $\omega_B$ | $\omega_C$ | $f_A$ | $f_B$ | $f_C$ |
|---|---|---|---|---|---|---|---|---|---|---|
| 235 | -0.06 | 3 | 1 | 10 | 4.95E-04 | 2.86E-04 | 1.81E-05 | 0.59 | 0.99 | 1.00 |
| 360 | -0.06 | 6 | 9 | 2 | 1.43E-04 | 9.41E-04 | 1.29E-04 | 0.61 | 0.99 | 0.68 |
| 68 | -0.05 | 1 | 3 | 5 | 2.27E-07 | 3.26E-05 | 8.31E-04 | 0.79 | 0.57 | 0.99 |
| 384 | -0.05 | 8 | 4 | 6 | 2.00E-07 | 7.32E-04 | 2.33E-05 | 0.62 | 1.00 | 0.99 |
| 328 | -0.04 | 5 | 5 | 7 | 1.26E-04 | 2.90E-06 | 5.84E-05 | 0.55 | 0.78 | 1.00 |
| 24 | -0.03 | 8 | 3 | 6 | 1.39E-07 | 5.17E-04 | 1.55E-04 | 0.74 | 0.94 | 1.00 |
| 106 | -0.03 | 2 | 10 | 5 | 5.43E-06 | 4.18E-06 | 4.75E-04 | 0.71 | 1.00 | 0.92 |
| 124 | -0.02 | 4 | 1 | 9 | 3.52E-07 | 2.24E-04 | 4.61E-04 | 0.54 | 0.58 | 0.98 |
| 355 | -0.02 | 8 | 7 | 4 | 2.16E-06 | 4.40E-05 | 3.99E-07 | 0.62 | 0.99 | 0.65 |
| 375 | -0.02 | 9 | 10 | 2 | 1.78E-07 | 1.62E-05 | 3.68E-04 | 0.58 | 0.90 | 1.00 |
| 380 | -0.02 | 8 | 4 | 6 | 7.99E-07 | 4.03E-04 | 4.40E-04 | 0.71 | 1.00 | 0.78 |
| 382 | -0.02 | 1 | 4 | 6 | 4.09E-05 | 2.53E-07 | 8.67E-07 | 0.70 | 1.00 | 0.69 |
| 413 | -0.02 | 8 | 1 | 9 | 1.29E-05 | 4.36E-07 | 6.13E-07 | 0.58 | 0.71 | 1.00 |
| 583 | -0.02 | 4 | 9 | 3 | 3.57E-07 | 1.14E-05 | 4.14E-04 | 0.69 | 0.74 | 0.99 |
| 875 | -0.02 | 5 | 3 | 7 | 1.85E-07 | 1.11E-07 | 2.30E-06 | 0.64 | 0.87 | 1.00 |
| 937 | -0.02 | 8 | 5 | 5 | 5.62E-05 | 1.56E-04 | 1.74E-04 | 0.78 | 0.84 | 0.98 |
| 79 | -0.01 | 7 | 5 | 1 | 1.12E-07 | 2.09E-04 | 5.39E-05 | 0.58 | 0.99 | 0.65 |
| 96 | -0.01 | 3 | 9 | 6 | 1.08E-04 | 9.15E-05 | 1.35E-06 | 0.80 | 0.95 | 0.99 |
| 164 | -0.01 | 8 | 6 | 1 | 1.93E-07 | 2.89E-07 | 9.11E-06 | 0.56 | 0.99 | 0.85 |
| 456 | -0.01 | 0 | 8 | 3 | 7.01E-04 | 6.16E-06 | 6.98E-05 | 0.55 | 0.99 | 0.83 |
| 591 | -0.01 | 1 | 1 | 8 | 4.35E-07 | 5.75E-06 | 1.05E-07 | 0.77 | 0.75 | 0.99 |
| 830 | -0.01 | 4 | 0 | 9 | 2.39E-05 | 1.93E-06 | 2.28E-06 | 0.81 | 0.76 | 0.98 |

**Appendix I: Model Equations - Deterministic:**

$$\frac{dS}{dt} = -\beta S\big((Y_S + Z_S) + F_A(Y_a + Z_a) + F_B(Y_b + Z_b) + \phi F_C(Y_c + Z_c) + F_{AB}(Y_{ab} + Z_{ab}) + F_{AC}(Y_{ac} + Z_{ac}) + F_{BC}(Y_{bc} + Z_{bc}) + F_{ABC}(Y_{abc} + Z_{abc})\big) + (1 - \xi_A\omega_A - \xi_B\omega_B - \xi_C\omega_C)T_S Y_S + \xi_B(1-\omega_B)T_S Y_a + \xi_C(1-\omega_C)T_S Y_a + \xi_A\kappa T_{SR} Y_a + \xi_C(1-\omega_C)Y_b + \xi_A(1-\omega_A)T_S Y_b + \xi_B\kappa T_{SR} Y_b + \xi_B(1-\omega_B)T_S Y_C + \xi_A(1-\omega_A)T_S Y_C + \xi_C\kappa T_{SR} Y_C + \xi_C(1-\omega_C)T_S Y_{ab} + (1-\xi_C)\kappa T_{SR} Y_{ab} + \xi_b(1-\omega_B)T_S Y_{ac} + (1-\xi_b)\kappa T_{SR} Y_{ac} + \xi_A(1-\omega_A)T_S Y_{bc} + (1-\xi_A)\kappa T_{SR} Y_{bc} + \kappa T_{SR} Y_{abc} + (1 - \xi_A\omega_A - \xi_B\omega_B - \xi_C\omega_C)T_m Z_S + \xi_B(1-\omega_B)T_m Z_a + \xi_C(1-\omega_C)T_m Z_a + \xi_A(1-\omega_A)T_m Z_b + \xi_C(1-\omega_C)T_m Z_b + \xi_B(1-\omega_B)T_m Z_c + \xi_A(1-\omega_A)T_m Z_C + \xi_C(1-\omega_C)T_m Z_{ab} + \xi_B(1-\omega_B)T_m Z_{ac} + \xi_A(1-\omega_A)T_m Z_{bc} + \delta(N - S)$$

$$\frac{dY_S}{dt} = \sigma\beta(Y_S + Z_S)S - \xi_A\omega_A T_S Y_S - \xi_B\omega_B T_S Y_S - \xi_C\omega_C T_S Y_S - (1 - \xi_A\omega_A - \xi_B\omega_B - \xi_C\omega_C)T_S Y_S - \delta Y_S$$

$$\frac{dY_a}{dt} = \sigma\beta F_A(Y_a + Z_a)S + \xi_A\omega_A T_S Y_S - \xi_B(1-\omega_B)T_S Y_a - \xi_C(1-\omega_C)T_S Y_a - \xi_B\omega_B T_S Y_a - \xi_C\omega_C T_S Y_a - \delta Y_a$$

$$\frac{dY_b}{dt} = \sigma\beta F_B(Y_b + Z_b)S + \xi_B\omega_B T_S Y_S - \xi_C(1-\omega_C)T_S Y_b - \xi_A(1-\omega_A)T_S Y_b - \xi_A\omega_A T_S Y_b - \xi_C\omega_C T_S Y_b - \delta Y_b$$

$$\frac{dY_c}{dt} = \sigma F_C\beta(Y_c + Z_c)S + \xi_C\omega_C T_S Y_S - \xi_B(1-\omega_B)T_S Y_c - \xi_A(1-\omega_A)T_S Y_c - \xi_A\omega_A T_S Y_c - \xi_B\omega_B T_S Y_c - \delta Y_c$$

$$\frac{dY_{ab}}{dt} = \sigma\beta F_{AB}(Y_{ab} + Z_{ab})S + \xi_B\omega_B T_S Y_a + \xi_A\omega_A T_S Y_b - \xi_C(1-\omega_C)T_S Y_{ab} - (1-\xi_C)\kappa T_{SR} Y_{ab} - \xi_C\omega_C T_S Y_{ab} - \delta Y_{ab}$$

$$\frac{dY_{ac}}{dt} = \sigma\beta F_{AC}(Y_{ac} + Z_{ac})S + \xi_C\omega_C T_S Y_a + \xi_A\omega_A T_S Y_c - \xi_B(1-\omega_B)T_S Y_{ac} - \xi_B\omega_B T_S Y_{ac} - (1-\xi_B)\kappa T_{SR} Y_{ac} - \delta Y_{ac}$$

$$\frac{dY_{bc}}{dt} = \sigma\beta F_{BC}(Y_{bc} + Z_{bc})S + \xi_C\omega_C T_S Y_b + \xi_B\omega_B T_S Y_c - \xi_A(1-\omega_A)T_S Y_{bc} - (1-\xi_A)\kappa T_{SR} Y_{bc} - \xi_A\omega_A T_S Y_{bc} - \delta Y_{bc}$$

$$\frac{dY_{abc}}{dt} = \sigma\beta F_{ABC}(Y_{abc} + Z_{abc})S + \xi_C\omega_C T_S Y_{ab} + \xi_B\omega_B T_S Y_{ac} + \xi_A\omega_A T_S Y_{bc} - \kappa T_{SR} Y_{abc} - \delta Y_{abc}$$

$$\frac{dZ_S}{dt} = (1-\sigma)\beta(Y_S + Z_S)S - \xi_A\omega_A T_M Z_S - \xi_B\omega_B T_M Z_S - \xi_C\omega_C T_M Z_S - (1 - \xi_A\omega_A - \xi_B\omega_B - \xi_C\omega_C)T_M Z_S - \delta Z_S$$

$$\frac{dZ_a}{dt} = (1-\sigma)\beta F_A(Y_a + Z_a)S + \xi_A\omega_A T_M Z_S - \xi_B(1-\omega_B)T_M Z_a - \xi_C(1-\omega_C)T_M Z_a$$

$$-\xi_B \omega_B T_M Z_a - \xi_C \omega_C T_M Z_a - \delta Z_a$$

$$\frac{dZ_b}{dt} = (1-\sigma)\beta F_B (Y_b + Z_b) S + \xi_B \omega_B T_M Z_S - \xi_C (1-\omega_C) T_M Z_b - \xi_A (1-\omega_A) T_M Z_b - \xi_C \omega_C T_M Z_b - \xi_A \omega_A T_M Z_b - \delta Z_b$$

$$\frac{dZ_c}{dt} = (1-\sigma) F_C \beta (Y_c + Z_c) S + \xi_C \omega_C T_M Z_S - \xi_B (1-\omega_B) T_M Z_c - \xi_A (1-\omega_A) T_M Z_C - \xi_A \omega_A T_M Z_C - \xi_B \omega_B T_M Z_C - \delta Z_C$$

$$\frac{dZ_{ab}}{dt} = (1-\sigma)\beta F_{AB} (Y_{ab} + Z_{ab}) S + \xi_B \omega_B T_M Z_a + \xi_A \omega_A T_M Z_b - \xi_C (1-\omega_C) T_M Z_{ab} - \xi_C \omega_C T_M Z_{ab} - \delta Z_{ab}$$

$$\frac{dZ_{ac}}{dt} = (1-\sigma)\beta F_{AC} (Y_{ac} + Z_{ac}) S + \xi_C \omega_C T_M Z_a + \xi_A \omega_A T_M Z_C - \xi_B (1-\omega_B) T_M Z_{ac} - \xi_B \omega_B T_M Z_{ac} - \delta Z_{ac}$$

$$\frac{dZ_{bc}}{dt} = (1-\sigma)\beta F_{BC} (Y_{bc} + Z_{bc}) S + \xi_C \omega_C T_M Z_b + \xi_B \omega_B T_M Z_c - \xi_A (1-\omega_A) T_M Z_{bc} - \xi_A \omega_A T_M Z_{bc} - \delta Z_{bc}$$

$$\frac{dZ_{abc}}{dt} = (1-\sigma)\beta F_{ABC} (Y_{abc} + Z_{abc}) S + \xi_c \omega_c T_M Z_{ab} + \xi_B \omega_B T_M Z_{ac} + \xi_A \omega_A T_M Z_{bc} - \delta Z_{abc}$$

### Appendix II: Model Equations – Stochastic

Recovery

| From | To | Rate |
|---|---|---|
| $(Y_S, S)$ | $(Y_S - 1, S + 1)$ | $((1 - \xi_A\omega_A - \xi_B\omega_B - \xi_C\omega_C)T_S + \delta)Y_S$ |
| $(Y_a, S)$ | $(Y_a - 1, S + 1)$ | $(\xi_B(1 - \omega_B)T_S + \xi_C(1 - \omega_C)T_S + \delta)Y_a$ |
| $(Y_b, S)$ | $(Y_b - 1, S + 1)$ | $(\xi_C(1 - \omega_C)T_S + \xi_A(1 - \omega_A)T_S + \delta)Y_b$ |
| $(Y_c, S)$ | $(Y_c - 1, S + 1)$ | $(\xi_B(1 - \omega_B)T_S + \xi_A(1 - \omega_A)T_S + \delta)Y_c$ |
| $(Y_{ab}, S)$ | $(Y_{ab} - 1, S + 1)$ | $(\xi_C(1 - \omega_C)T_S + (1 - \xi_C)\kappa T_{SR} + \delta)\, Y_{ab}$ |
| $(Y_{ac}, S)$ | $(Y_{ac} - 1, S + 1)$ | $(\xi_B(1 - \omega_B)T_S + (1 - \xi_B)\kappa T_{SR} + \delta)\, Y_{ac}$ |
| $(Y_{bc}, S)$ | $(Y_{bc} - 1, S + 1)$ | $(\xi_A(1 - \omega_A)T_S + (1 - \xi_A)\kappa T_{SR} + \delta)\, Y_{bc}$ |
| $(Y_{abc}, S)$ | $(Y_{abc} - 1, S + 1)$ | $(\kappa T_{SR} + \delta)Y_{abc}$ |
| $(Z_S, S)$ | $(Z_S - 1, S + 1)$ | $((1 - \xi_A\omega_A - \xi_B\omega_B - \xi_C\omega_C)T_M + \delta)Z_S$ |
| $(Z_a, S)$ | $(Z_a - 1, S + 1)$ | $(\xi_B(1 - \omega_B)T_M + \xi_C(1 - \omega_C)T_M + \delta)Z_a$ |
| $(Z_b, S)$ | $(Z_b - 1, S + 1)$ | $(\xi_C(1 - \omega_C)T_M + \xi_A(1 - \omega_A)T_M + \delta)Z_b$ |
| $(Z_c, S)$ | $(Z_c - 1, S + 1)$ | $(\xi_B(1 - \omega_B)T_M + \xi_A(1 - \omega_A)T_M + \delta)Z_c$ |
| $(Z_{ab}, S)$ | $(Z_{ab} - 1, S + 1)$ | $(\xi_C(1 - \omega_C)T_S + \delta)\, Z_{ab}$ |
| $(Z_{ac}, S)$ | $(Z_{ac} - 1, S + 1)$ | $(\xi_B(1 - \omega_B)T_M + \delta)\, Z_{ac}$ |
| $(Z_{bc}, S)$ | $(Z_{bc} - 1, S + 1)$ | $(\xi_A(1 - \omega_A)T_M + \delta)\, Z_{bc}$ |
| $(Z_{abc}, S)$ | $(Z_{abc} - 1, S + 1)$ | $\delta Z_{abc}$ |

Infection

| From | To | Rate |
|---|---|---|
| $(S\ , Y_S)$ | $(S-1, Y_S+1)$ | $\sigma\beta(Y_S+Z_S)S$ |
| $(S\ , Y_a)$ | $(S-1, Y_a+1)$ | $\sigma\beta F_A(Y_a+Z_a)S$ |
| $(S\ , Y_b)$ | $(S-1, Y_b+1)$ | $\sigma\beta F_B(Y_b+Z_b)S$ |
| $(S\ , Y_c)$ | $(S-1, Y_c+1)$ | $\sigma F_C\beta(Y_c+Z_c)S$ |
| $(S\ , Y_{ab})$ | $(S-1, Y_{ab}+1)$ | $\sigma\beta F_{AB}(Y_{ab}+Z_{ab})S$ |
| $(S\ , Y_{ac})$ | $(S-1, Y_{ac}+1)$ | $\sigma\beta F_{AC}(Y_{ac}+Z_{ac})S$ |
| $(S\ , Y_{bc})$ | $(S-1, Y_{bc}+1)$ | $\sigma\beta F_{BC}(Y_{bc}+Z_{bc})S$ |
| $(S\ , Y_{abc})$ | $(S-1, Y_{abc}+1)$ | $\sigma\beta F_{ABC}(Y_{abc}+Z_{abc})S$ |
| $(S\ , Z_S)$ | $(S-1, Z_S+1)$ | $(1-\sigma)\beta(Y_S+Z_S)S$ |
| $(S\ , Z_a)$ | $(S-1, Z_a+1)$ | $(1-\sigma)\beta F_A(Y_a+Z_a)S$ |
| $(S\ , Z_b)$ | $(S-1, Z_b+1)$ | $(1-\sigma)\beta F_B(Y_b+Z_b)S$ |
| $(S\ , Z_c)$ | $(S-1, Z_c+1)$ | $(1-\sigma)F_C\beta(Y_c+Z_c)S$ |
| $(S\ , Z_{ab})$ | $(S-1, Z_{ab}+1)$ | $(1-\sigma)\beta F_{AB}(Y_{ab}+Z_{ab})S$ |
| $(S\ , Z_{ac})$ | $(S-1, Z_{ac}+1)$ | $(1-\sigma)\beta F_{AC}(Y_{ac}+Z_{ac})S$ |
| $(S\ , Z_{bc})$ | $(S-1, Z_{bc}+1)$ | $(1-\sigma)\beta F_{BC}(Y_{bc}+Z_{bc})S$ |
| $(S\ , Z_{abc})$ | $(S-1, Z_{abc}+1)$ | $(1-\sigma)\beta F_{ABC}(Y_{abc}+Z_{abc})S$ |

Resistance Evolution

| From | To | Rate |
|---|---|---|
| $(Y_S, Y_a)$ | $(Y_S - 1, Y_a + 1)$ | $\xi_A \omega_A T_S Y_S$ |
| $(Y_S, Y_b)$ | $(Y_S - 1, Y_b + 1)$ | $\xi_B \omega_B T_S Y_S$ |
| $(Y_S, Y_c)$ | $(Y_S - 1, Y_c + 1)$ | $\xi_C \omega_C T_S Y_S$ |
| $(Y_a, Y_{ab})$ | $(Y_a - 1, Y_{ab} + 1)$ | $\xi_B \omega_B T_S Y_a$ |
| $(Y_a, Y_{ac})$ | $(Y_a - 1, Y_{ac} + 1)$ | $\xi_C \omega_C T_S Y_a$ |
| $(Y_b, Y_{ab})$ | $(Y_b - 1, Y_{ab} + 1)$ | $\xi_A \omega_A T_S Y_b$ |
| $(Y_b, Y_{bc})$ | $(Y_b - 1, Y_{bc} + 1)$ | $\xi_C \omega_C T_S Y_b$ |
| $(Y_c, Y_{ac})$ | $(Y_c - 1, Y_{ac} + 1)$ | $\xi_A \omega_A T_S Y_c$ |
| $(Y_c, Y_{bc})$ | $(Y_c - 1, Y_{bc} + 1)$ | $\xi_B \omega_B T_S Y_c$ |
| $(Y_{ab}, Y_{abc})$ | $(Y_{ab} - 1, Y_{abc} + 1)$ | $\xi_C \omega_C T_S Y_{ab}$ |
| $(Y_{ac}, Y_{abc})$ | $(Y_{ac} - 1, Y_{abc} + 1)$ | $\xi_B \omega_B T_S Y_{ac}$ |
| $(Y_{bc}, Y_{abc})$ | $(Y_{bc} - 1, Y_{abc} + 1)$ | $\xi_A \omega_A T_S Y_{bc}$ |
| $(Z_S, Z_a)$ | $(Z_S - 1, Z_a + 1)$ | $\xi_A \omega_A T_M Z_S$ |
| $(Z_S, Z_b)$ | $(Z_S - 1, Z_b + 1)$ | $\xi_B \omega_B T_M Z_S$ |
| $(Z_S, Z_c)$ | $(Z_S - 1, Z_c + 1)$ | $\xi_C \omega_C T_M Z_S$ |
| $(Z_a, Z_{ab})$ | $(Z_a - 1, Z_{ab} + 1)$ | $\xi_B \omega_B T_M Z_a$ |
| $(Z_a, Z_{ac})$ | $(Z_a - 1, Z_{ac} + 1)$ | $\xi_C \omega_C T_M Z_a$ |
| $(Z_b, Z_{ab})$ | $(Z_b - 1, Z_{ab} + 1)$ | $\xi_A \omega_A T_M Z_b$ |
| $(Z_b, Z_{bc})$ | $(Z_b - 1, Z_{bc} + 1)$ | $\xi_C \omega_C T_M Z_b$ |
| $(Z_c, Z_{ac})$ | $(Z_c - 1, Z_{ac} + 1)$ | $\xi_A \omega_A T_M Z_C$ |
| $(Z_c, Z_{bc})$ | $(Z_c - 1, Z_{bc} + 1)$ | $\xi_B \omega_B T_M Z_C$ |
| $(Z_{ab}, Z_{abc})$ | $(Z_{ab} - 1, Z_{abc} + 1)$ | $\xi_C \omega_C T_M Z_{ab}$ |
| $(Z_{ac}, Z_{abc})$ | $(Z_{ac} - 1, Z_{abc} + 1)$ | $\xi_B \omega_B T_M Z_{ac}$ |
| $(Z_{bc}, Z_{abc})$ | $(Z_{bc} - 1, Z_{abc} + 1)$ | $\xi_A \omega_A T_M Z_{bc}$ |
